# Interscapular fat is associated with impaired glucose tolerance and insulin resistance independent of visceral fat mass

**DOI:** 10.1101/2022.02.07.22270571

**Authors:** Andreas Vosseler, Jürgen Machann, Louise Fritsche, Christian Kübler, Hans-Ulrich Häring, Andreas L. Birkenfeld, Norbert Stefan, Andreas Peter, Andreas Fritsche, Robert Wagner, Martin Heni

## Abstract

Dysregulated body fat distribution is a major determinant of various diseases. Adipose tissue in different localizations of the body appear to have protective or detrimental properties. Particularly increased visceral fat mass and ectopic lipids in the liver are linked to metabolic disorders such as insulin resistance and type 2 diabetes. Furthermore, interscapular fat is considered to be a non-classical, but metabolically active, fat compartment.

In this study, we measured interscapular fat mass and investigated its relationship with glucose tolerance, insulin sensitivity and insulin secretion in 822 subjects with a wide range of body mass index (BMI) and different glucose tolerance status. Magnetic resonance imaging was used to quantify body fat depots and an oral glucose tolerance test (OGTT) was performed to determine glucose metabolism.

Elevated interscapular fat mass was positively associated with age, BMI, total body, visceral and subcutaneous adipose tissue mass. High interscapular fat mass associated with elevated fasting glucose levels, glucose levels at 2 hours during the OGTT, glycated hemoglobin, as well as with insulin resistance, independently of sex, age, total body and visceral fat mass.

In conclusion, interscapular fat might be a highly specific fat compartment with potential impact on glucose metabolism and the pathogenesis of diabetes mellitus. Since this depot is assessible by ultrasound, it could represent a feasible target to quantify metabolic risk in the future.

## Introduction

There are large inter-individual differences in the distribution of fat across the body. This distribution of adipose tissue appears to be crucial for human health as it is believed to contribute to the pathogenesis of various diseases^1,2^. Body fat distribution partly depends on sex and age^1,2^. The majority of body fat (around 80% of total body fat) is stored as subcutaneous adipose tissue (SCAT), most prominently around the abdomen as well as in the subscapular, gluteal and femoral areas^3^, with potentially distinct impact dependent on its location^4,5^.

The second major fat compartment, visceral adipose tissue (VAT), accounts for up to 20 % of total body fat content^6^. In contrast to subcutaneous fat, the venous blood of visceral adipose tissue is drained directly towards the liver via the portal vein^6^. Factors released from this fat compartment activate immune functions both directly in adipose tissue and in the liver. In response, cytokines are released with unfavorable impact on the entire body e.g. vascular inflammation^6,7^ that increases the risk for cardiovascular diseases^8,9^. Furthermore, visceral fat is associated with multiple complications such as hypertension, dyslipidemia, insulin resistance and type 2 diabetes^10,11^. Visceral fat is also a predictor of all-cause mortality^12,13^.

In addition to increased visceral fat mass, other fat compartments might also have a negative impact on glucose and lipid metabolism. Particularly fat accumulation in the liver and fat that is localized in the neck area, are related to insulin resistance and cardiovascular diseases in humans ^14–20^.

Besides different fat locations, there are functionally and histologically different types of adipocytes that are categorized as white adipose tissue (WAT) and brown adipose tissue (BAT), as well as brown-in-white, so called beige fat^21,22^.

Though, BAT accounts only for a small proportion of adipose tissue^23^. The two fat types appear to have antagonistic characteristics as white adipose tissue stores excess energy and brown adipose tissue primarily converts stored energy to heat^24^. While brown adipose tissue substantially contributes to the total amount of fat in rodents, the proportion of brown adipose tissue is much smaller in humans^25^. This type of fat is more prominent in newborns but can be activated by cold exposure also in adults^26^ where it inversely correlates with BMI^27^ and glucose metabolism^28,29^. In infants, brown adipose tissue is present in the interscapular region^30^, while in adults it appears to be mainly located elsewhere^31^.

In contrast to theoretical beneficial characteristics of brown adipose tissue in terms of glucose metabolism, we previously detected an association of increased interscapular fat mass with insulin resistance in 168 subjects^17^. In line, the neck region in whole-body MR images was highlighted in over half of the cases by machine learning as important to detect diabetes in a recent study^32^. In contrast to visceral adipose tissue, that can be precisely quantified only by expensive approaches or approaches that relay on radiation, interscapular fat is assessable by easier approaches like ultrasonography^33,34^.

In the present study, we extended our previous work^17^ and reevaluated this non-classical fat depot to study possible links of interscapular fat with glucose metabolism in a much larger cohort.

## Methods

In our study we could now include 822 volunteers, 510 females and 312 males. The study participants were recruited from the ongoing TUEFF-study between 2011 and 2020^32^. All volunteers gave written informed consent and the study was approved by the Ethics Committee of the University Hospital of Tübingen, Germany, and all research was conducted in accordance with relevant guidelines and regulations.

The volunteers had an increased risk for the development of type 2 diabetes due to at least one risk factor (BMI >27 kg/m^2^ or obesity, impaired fasting glucose or glucose tolerance, previous gestational diabetes or type 2 diabetes in first-grade relatives)^35^. An oral glucose tolerance test (OGTT) was performed after 10 hours of overnight fast. All volunteers underwent the OGTT with 75 g dextrose (Accu-Chek Dextrose OGT, Roche). Blood samples were obtained before and 30, 60, 90 and 120 minutes after glucose ingestion.

Baseline blood samples included glycohemoglobin A1c (HbA1c) that was measured with Tosoh glycohemoglobin analyzer HLC-723G8 (Tosoh Bioscience, Tokyo, Japan).

Pro-insulin, insulin, C-peptide and cortisol levels were determined by ADVIA Centaur XP Immunoassay System (Siemens Healthineers, Eschborn, Germany). Glucose measurements were analyzed by hexokinase method with ADVIA XPT System (Siemens Healthineers, Eschborn, Germany). All measurements were performed in a routine diagnostic laboratory accredited with the German accredited body (DAkkS). Insulin sensitivity was estimated from oral glucose tolerance tests as described by Matsuda and DeFronzo^36^ (ISI Matsuda). NEFA-based insulin sensitivity index (NEFA-ISI) was calculated including BMI, serum insulin and non-esterified fatty acids (NEFA) as proposed by Wagner et al.^37^. First phase insulin secretion was calculated as proposed by Stumvoll et al.^38^: 1283 + 1.829 x Insulin_30_ – 138.7 x Glucose_30_ + 3.772 x Insulin_0._

Body fat compartments and liver fat content were quantified by magnetic resonance imaging (MRI) and proton magnetic resonance spectroscopy (^1^H-MRS), respectively^39^. In brief, a T1-weighted fast spin echo sequence was applied to acquire images from the whole body in a total measurement time of approximately 20 minutes. From these, visceral adipose tissue (between hips and thoracic diaphragm) and subcutaneous adipose tissue (between hips and shoulders) were quantified using an automated fuzzy c-means algorithm and orthonormal snakes^40^. Additionally, interscapular fat area in the neck was segmented between the humeral heads as shown in figure 1. Liver fat was assessed in the posterior part of segment 7 applying a single voxel STEAM technique and calculating the ratio from fat (methylene + methyl signals) and the sum of water and fat resonances.

**Figure 1:**
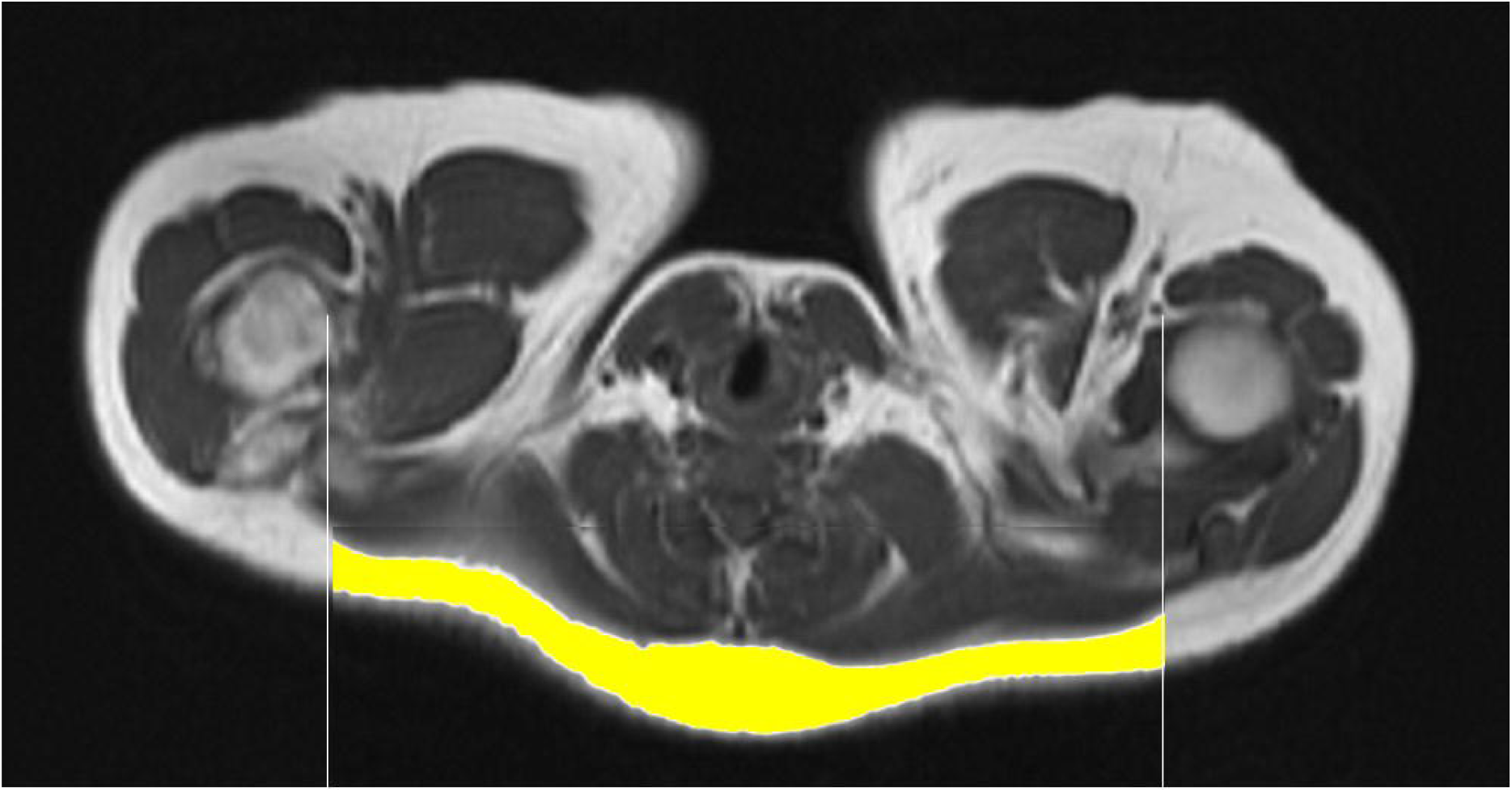
Interscapular fat depot. Magnetic resonance image of the interscapular fat depot, area in the neck was segmented between the humeral heads (highlighted section).

Statistical analysis on log_*e*_-transformed data was performed using JMP 14 (SAS Institute, Cary, NC). Two-sided t-tests were performed to compare variables. Data were furthermore tested with multivariate models. The data are presented as mean ± SE, p < 0.05 is considered statistically significant. Regression coefficient is presented as standardized β.

## Results

The average age of the participants was 46 years (SD ± 15) and the body mass index (BMI) was 29.4 kg/m2 (SD ± 6.3). The basic anthropometrics are displayed in table 1. 50.9% of the volunteers (n=418) had a normal glucose tolerance (neither impaired fasting glucose nor impaired glucose tolerance), 24.9% (n=205) had an impaired fasting glucose (IFG, fasting plasma glucose levels 5.6 mmol/l to 6.9 mmol/l), 96 were categorized as impaired glucose tolerance (IGT, 2-h values in the oral glucose tolerance test of 7.8 mmol/l to 11.0 mmol/l) and 103 had both IFG and IGT, according to ADA’s criteria^41^. The metabolic characterization of our cohort is shown in table 2.

Interscapular fat mass was higher in females than in males, both unadjusted (β=0.07±0.629; p=0.04) and adjusted for total adipose tissue mass (β=-0.07±0.002, p=0.002). However, after additional adjustment for visceral fat mass, sex was no longer associated with interscapular fat (β=-0.05±0.503; p=0.10). Interscapular fat was positively associated with age (β=0.12±0.029; p=0.0005). Furthermore, it was positively correlated with BMI (β=0.82±0.007; p<0.0001), total adipose tissue (β=0.82±0.017; p<0.0001), visceral adipose tissue (β=0.53±0.004; p<0.0001) and with subcutaneous adipose tissue (β=0.84±0.007; p<0.0001), detailed results are displayed in table 3 and figure 2.

**Figure 2:**
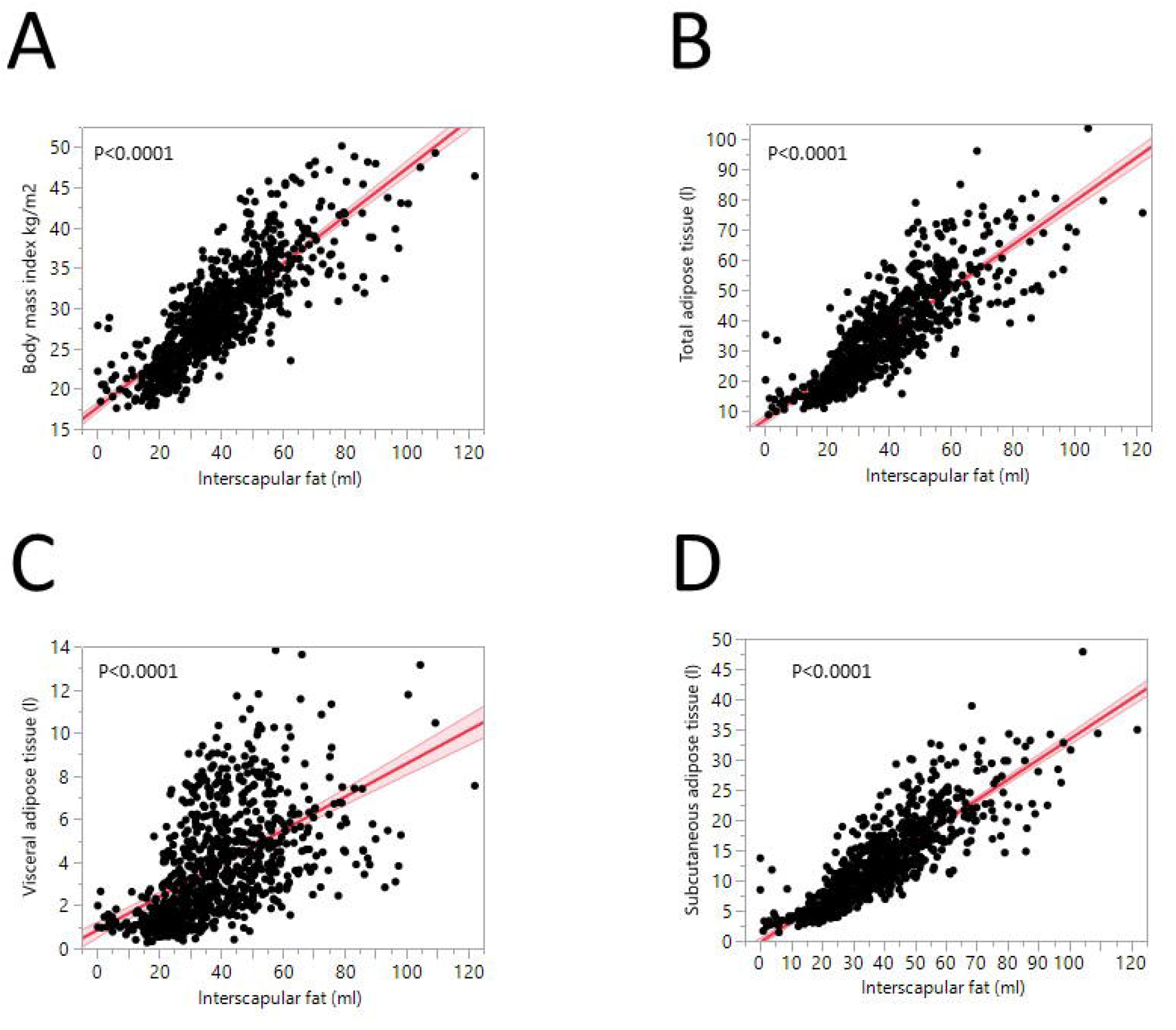
Correlation of interscapular fat with body mass index and fat depots. Positive correlation of interscapular fat with body mass index (A), total adipose tissue (B), visceral adipose tissue (C) and subcutaneous adipose tissue (D) in a univariate model, n=822. Lines represent fit lines ±CI.

We next analyzed associations of interscapular fat with glucose metabolism (table 4). This fat compartment was positively correlated with fasting glucose (β=0.26±0.002; p<0.0001). This relationship remained significant after adjustment for sex and total adipose tissue (β=0.14±0.002; p=0.008), as well as after additional adjustment visceral adipose tissue (β=0.12±0.002; p=0.02). Of note, both interscapular and visceral fat were independently associated with fasting glucose (β=0.23±0.015; p=0.0002).

Interscapular fat was also associated with 2 hours glucose levels during the OGTT (β=0.24±0.003; p<0.0001). Again, this relationship was independent of sex, age, total and visceral fat content (β=0.24±0.005; p<0.0001). In line, there were positive associations of interscapular fat with AUC_glucose_ during the OGTT (β=0.30±0.006; p<0.0001;) as well as with HbA1c (β=0.21±0.009; p<0.0001) in univariate models. These associations remained statistically significant after adjustment for sex, age, total and visceral fat content (AUC_glucose_ during the OGTT: β=0.29±0.010; p<0.0001; HbA1c: β=0.13±0.013; p=0.02).

In search for potential contributors to the relation of interscapular fat and glycemia, we next analyzed the two major determinants of blood glucose – insulin sensitivity and insulin secretion. Interscapular fat content was negatively associated with insulin sensitivity (ISI Matsuda: β=-0.51±0.013; p<0.0001; NEFA ISI: β=-0.66±0.003; p<0.0001) as shown in figure 3. This association remained significant after adjustment for sex, age and total adipose tissue mass (ISI Matsuda: β=-0.27±0.023; p<0.0001; NEFA ISI: β=-0.30±0.005; p<0.0001) as well as after additional adjustment visceral adipose tissue content (ISI Matsuda: β=-0.22±0.02; p<0.0001; NEFA ISI: β=-0.27±0.005; p<0.0001).

**Figure 3:**
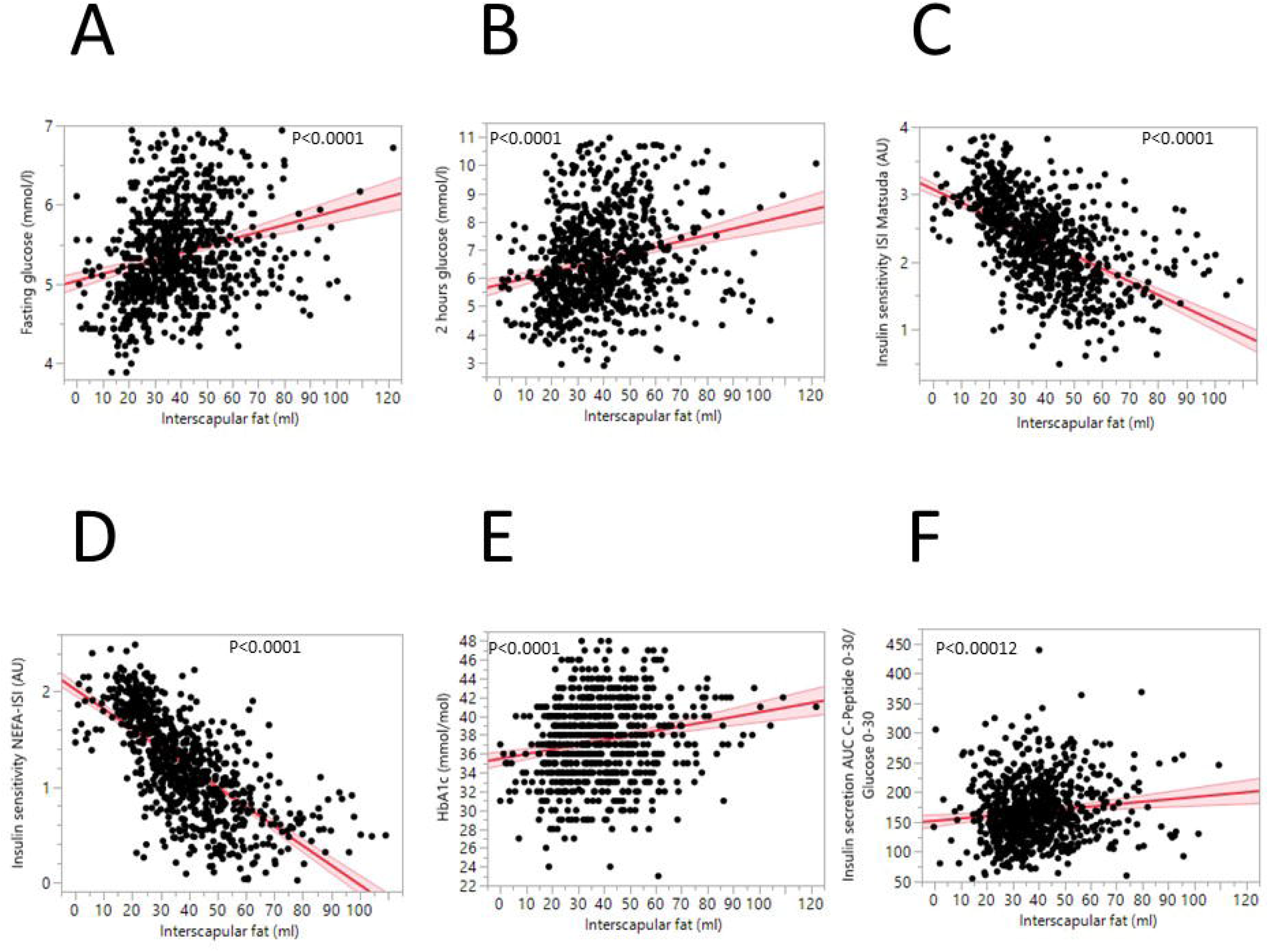
Correlation of interscapular fat with parameters of glucose metabolism. Positive correlation of interscapular fat with fasting glucose (A), 2 hours glucose from the oral glucose tolerance test (B), insulin sensitivity index ISI Matsuda (C), insulin sensitivity index NEFA ISI (D) and glycated hemoglobin HbA1c in an univariate model, n=822. Lines represent fit lines ±CI.

Only in a univariate model, insulin secretion assessed from both C-peptide and glucose concentrations was positively associated with interscapular fat (AUC_C_-_pep0-30/AUCgluc0-30:_ β=0.11±0.123; p=0.001; adjusted for ISI Matsuda). However, after adjustment for sex, age, total and visceral adipose tissue mass, there was no significant association of insulin secretion with interscapular fat (AUC_C_-_pep0-30/AUCgluc0-30:_ β=-0.04±0.187; p=0.45; adjusted for ISI Matsuda).

First phase insulin secretion assessed from both serum insulin and glucose concentrations was positively correlated with interscapular fat again only in univariate models (β=0.16±1.458; p<0.0001; adjusted for ISI Matsuda). However, there was no correlation of first phase insulin secretion with interscapular fat after adjustment for sex, age, total and visceral adipose tissue content (β=-0.03±2.171; p=0.61; adjusted for ISI Matsuda).

There were no significant interactions of interscapular fat with serum cortisol (β=-0.06±0.321; p=0.11).

## Discussion

We detected an association of higher amounts of interscapular fat with impaired glucose tolerance and insulin resistance. In addition, the link between interscapular fat and glycemia was robust and independent of the major potential confounders sex, age and total adiposity. Of note, the detected associations were independent of visceral fat mass. Though, there was no correlation of interscapular fat with insulin secretion in our multivariable models. The fact that the detected associations with glycemia remained significant after adjustment for visceral fat suggests that interscapular fat is not just a read-out of visceral fat mass.

Our data support the earlier hypothesis^17^ that interscapular fat is linked to insulin resistance in humans. Thus, interscapular fat tissue might harbor cells that are metabolically active and, thereby, contribute to the pathogenesis of insulin resistance in the entire body. The negative effects of interscapular fat on glucose metabolism could be caused by pro-inflammatory adipokines, similarly to what has been described for other non-classical fat compartments, like perivascular fat^42^.

The main driver of the development of detrimental fat depots such as interscapular fat is obesity that also induces excess fat depots that lead to insulin resistance via cellular mechanisms^43^.

In addition, elevated cortisol levels can contribute to insulin resistance^44^. Especially fat in the neck area is a symptom of pathological hypercortisolism and might therefore be a relevant parameter in accordance with interscapular fat. Though, interscapular fat was not associated with serum cortisol in our study. Since we measured cortisol only once in the morning, it is still possible that one pathophysiologic link could be subclinical hypercortisolism that would only be detectable by more measurements over the day or by specific suppression tests^45,46^. The association of interscapular fat and insulin resistance in our study was consistent with previous findings with MR-derived quantification^17^ or simpler anthropometric estimates^47,48^. In line, interscapular fat was associated with additional risk factors for insulin resistance like age, increased BMI and body fat compartments.

While the amount of interscapular fat was higher in women, this sex difference was not independent of total and visceral fat content. This observation is well in line with known differences in body fat distribution between sexes^3^ but argues against a sex-specific effect specifically for interscapular fat.

Interscapular fat might be a clinically relevant risk factor of impaired glucose tolerance, independent of visceral fat mass, that is in turn associated with an increased risk of cardiovascular disease^49,50^.

Furthermore, insulin secretion was not linked to interscapular fat after adjustment for potential confounders.

In line with previous observation in very large cohorts^31^, our data argue against a predominance of highly active brown fat in the interscapular region as protective properties of this fat depot on glucose metabolism are missing. In addition, the association of interscapular fat with insulin resistance has an opposing direction to the characteristics of brown fat in other studies^51,52^.

Even though it appears unlikely that interscapular fat represents larger amounts of brown adipose tissue, a limitation of our study is that the histological type of the adipose tissue of interscapular fat was not examined. In addition, potential risk factors for the accumulation of interscapular fat stay unclear.

In conclusion, interscapular fat is associated with insulin resistance and fasting glucose as well as 2-hours glucose from the OGTT. It seems to be an important fat compartment concerning glucose metabolism, independent of visceral fat mass. Further studies may elucidate its consequence for the pathogenesis of insulin resistance and dysglycemia. Since this compartment is assessable by widely available approaches like ultrasound^33,34^, it could represent a feasible target to quantify metabolic risk in clinical practice in the future.

## Data Availability

The data are not publicly available due to them containing information that could compromise research participant privacy/consent.

## Author Contributions

AV researched and analyzed data and drafted the manuscript. JM researched and analyzed data and contributed to discussion. LF, CK, AP, NS, AF and RW researched data and contributed to discussion. HUH and AB contributed to discussion. MH contributed to analyses, supervised the project and contributed to discussion. All authors approved the final version of the manuscript prior to submission.

## Acknowledgements

We thank the research volunteers for participating in the study. We gratefully acknowledge the excellent technical assistance of K. Waneck, A. Eberle, I. Wagener, E. Stehle, E. Metzinger, D. Neuscheler (all University of Tübingen).

