## Supplementary material for "Interscapular fat is associated with impaired glucose tolerance and insulin resistance independent of visceral fat mass": Table 1

**Table 1. Basic anthropometric**

| n | 822 |
| --- | --- |
| Sex | females 510; males 312 |
| Age (years) | 46 (±15) |
| BMI (kg/m^2^) | 29.4 (±6.3) |
| Waist circumference (cm) | 95.1 (±15.8) |
| Hip circumference (cm) | 106.5 (±13.3) |
| Waits-to-hip-ratio | 0.89 (±0.09) |
| Body fat content (%, BIA-derived) | 34.1 (±12.3) |

**values are given as mean ±SD**
