## Supplementary material for "Interscapular fat is associated with impaired glucose tolerance and insulin resistance independent of visceral fat mass": Table 2

**Table 2. Metabolic characteristics: Glucose status, insulin sensitivity indices and body fat distribution.**

| Fasting glucose (mmol/l) | 5.38 (±0.61) n=821 |
| --- | --- |
| Glucose 120 min (mmol/l) | 6.62 (±1.64) n=820 |
| AUC_Glucose_ during OGTT | 15.56 (±3.38) n=818 |
| AUC_C_-_pep0-30/AUCgluc0-30_ | 167.55 (±60.24) n=818 |
| HbA1c (mmol/mol / %) | 37.39 (±4.24) / 5.57 (±0.39) n=751 |
| Normal glucose tolerance (NGT) | n=418 (50.9%) |
| Impaired fasting glucose (IFT) | n=205 (24.9%) |
| Impaired glucose tolerance (IGT) | n=96 (11.7%) |
| Both IFG and IGT | n=103 (12.5%) |
| Insulin sensitivity index, OGTT-derived, Matsuda (AU) | 12.25 (±7.82) n=812 |
| NEFA insulin sensitivity index, OGTT-derived (AU) | 3.89 (±2.04) n=776 |
| Total adipose tissue (l) | 35.6 (± 15.4) n=817 |
| Visceral adipose tissue (l) | 4.0 (± 2.5) n=819 |
| Subcutaneous adipose tissue (l) | 13.0 (± 7.0) n=819 |
| Liver fat content (%) | 6.3 (±6.6) |
| Interscapular fat content (ml) | 39.0 (±17.5) |

**values are given as mean ±SD**
