## Supplementary material for "Interscapular fat is associated with impaired glucose tolerance and insulin resistance independent of visceral fat mass": Table 3

**Table 3. Univariate models testing associations of interscapular fat with sex, age, BMI, total adipose tissue (TAT), visceral adipose tissue (VAT), subcutaneous adipose tissue (SCAT) and cortisol.**

| **Interscapular fat (IF)** |  | **p** | **Standardized β** | **Standard error SE** |
| --- | --- | --- | --- | --- |
|  | Sex | p=0.04 | β=0.07 | 0.629 |
|  | Age | p=0.0005 | β=0.12 | 0.029 |
|  | BMI | p<0.0001 | β=0.82 | 0.007 |
|  | TAT/l | p<0.0001 | β=0.82 | 0.017 |
|  | VAT/l | p<0.0001 | β=0.53 | 0.004 |
|  | SCAT/l | p<0.0001 | β=0.84 | 0.007 |
|  | Cortisol | p=0.11 | β=-0.06 | 0.321 |

**values are given as mean ±SE; n=822**
