## Supplementary material for "Interscapular fat is associated with impaired glucose tolerance and insulin resistance independent of visceral fat mass": Table 4

**Table 4. Multivariate models testing associations of interscapular fat with glucose metabolism. Adjusted for sex, age, total and visceral adipose tissue mass.**

|  | **p** | **Standardized β** | **Standard error SE** |
| --- | --- | --- | --- |
| **AUC _Glucose 0-120_** | p<0.0001 | β=0.29 | 0.010 |
| **Glucose 120** | p<0.0001 | β=0.24 | 0.005 |
| **HbA1c** | p=0.02 | β=0.13 | 0.013 |
| **Glucose 0** | p=0.02 | β=0.12 | 0.002 |
| **First phase insulin secretion** | p=0.61 | β=-0.03 | 2.171 |
| **AUC _C-peptide 0-30_ / AUC _Glucose 0-30_** | p=0.45 | β=-0.04 | 0.187 |
| **ISI Matsuda** | p<0.0001 | β=-0.22 | 0.022 |
| **NEFA ISI** | p<0.0001 | β=-0.27 | 0.005 |

**values are given as mean ±SE; n=819**
